# Correcting B0 inhomogeneity-induced distortions in whole-body diffusion MRI of bone

**DOI:** 10.1101/2020.11.22.20236547

**Authors:** Leonardino A. Digma, Christine H. Feng, Christopher C. Conlin, Ana E. Rodríguez-Soto, Allison Y. Zhong, Troy S. Hussain, Asona Lui, Kanha Batra, Aaron B. Simon, Roshan Karunamuni, Joshua Kuperman, Rebecca Rakow-Penner, Michael E. Hahn, Anders M. Dale, Tyler M. Seibert

## Abstract

**Objectives:** Diffusion-weighted magnetic resonance imaging (DWI) of the musculoskeletal system has various applications, including visualization of bone tumors. However, DWI acquired with echo-planar imaging is susceptible to distortions due to static magnetic field inhomogeneities. This study aimed to estimate spatial displacements of bone and to examine whether distortion corrected DWI images more accurately reflect underlying anatomy.

**Methods:** Whole-body MRI data from 127 prostate cancer patients were analyzed. The reverse polarity gradient (RPG) technique was applied to DWI data to estimate voxel-level distortions and to produce a distortion corrected DWI dataset. First, an anatomic landmark analysis was conducted, in which corresponding vertebral landmarks on DWI and anatomic *T*_*2*_-weighted images were annotated. Changes in distance between DWI- and *T*_*2*_-defined landmarks (i.e., changes in error) after distortion correction were calculated. In secondary analyses, distortion estimates from RPG were used to assess spatial displacements of bone metastases. Lastly, changes in mutual information between DWI and *T*_*2*_-weighted images of bone metastases after distortion correction were calculated.

**Results:** Distortion correction reduced anatomic error of vertebral DWI up to 29 mm. Error reductions were consistent across subjects (Wilcoxon signed-rank p<10^−20^). On average (±SD), participants’ largest error reduction was 11.8 mm (±3.6). Mean (95% CI) displacement of bone lesions was 6.0 mm (95% CI: 5.1-7.0); maximum displacement was 17.1 mm. Corrected diffusion images were more similar to structural MRI, as evidenced by consistent increases in mutual information (Wilcoxon signed-rank p<10^−12^).

**Discussion:** These findings support the use of distortion correction techniques to improve localization of bone on DWI.

**Key Points:**

- Diffusion weighted images of bone tissue undergo substantial spatial distortions when acquired with echo-planar imaging.
- These distortions can be efficiently corrected with the reverse polarity gradient technique to generate diffusion images that more accurately reflect underlying anatomy.
- In the context of bone tumor imaging where precise localization may be required, distortion correction techniques, such as reverse polarity gradient, should be applied.

## Introduction

Diffusion-weighted magnetic resonance imaging (DWI) of the musculoskeletal system has wide-ranging applications, such as visualization of vertebral fractures, ligament tears, bone tumors, and assessment of bone quality with aging and osteoporosis [1, 2]. In each of these applications, DWI offers several advantages. In the context of bone tumors, for example, DWI is highly sensitive and has improved spatial resolution compared to the more commonly used technetium-99m skeletal scintigraphy. Furthermore, DWI does not expose patients to radiation, unlike bone scan, positron emission tomography, and computed tomography (PET/CT) scan, which are also frequently used to evaluate metastases.

Despite its advantages, DWI of bone suffers from technical limitations that limit its clinical adoption. One limitation is distortion induced by static magnetic field (*B*_*0*_) inhomogeneity. This distortion is inevitably present in echo-planar imaging techniques used to collect DWI [3] and can cause substantial warping of images. Several techniques have been developed to measure and correct for these artifacts [4–6], but none has yet been widely adopted for routine clinical use. For DWI of prostate and breast tissue, distortion correction using the reverse polarity gradient (RPG) technique [5] has been shown to improve anatomic localization [7, 8]. The impact of these distortions has not yet been explored in the context of bone or whole-body DWI acquisitions.

In this study, we examined *B*_*0*_ inhomogeneities in whole-body DWI and investigated how they can affect visualization of bone anatomy and localization of bone metastases. We used RPG to estimate the magnitude of these distortions in images of bone and to determine whether correcting for these distortions would improve anatomic correlation of skeletal DWI with *T*_*2*_-weighted images. We also performed a secondary analysis to explore how these distortions affect localization of bone metastases.

## Methods

### Study Population

Patients with suspected or known metastatic prostate cancer were enrolled in a prospective, observational, non-contrast whole-body MRI trial from August 2017 to October 2020. All study participants provided informed consent.

### Image Acquisition and Processing

Whole-body MRI was acquired on a 3.0 T MRI system (Discovery MR750, GE Healthcare, Waukesha, WI, USA) with a multi-parametric MRI protocol that included *T*_*2*_-weighted (TE/TR: 113/1350 ms, acquisition matrix: 384×224, resampled matrix: 512×512, FOV: 400 x 400 mm, slices: 46, slice thickness: 6 mm) and DWI (TE/TR: 75/4750 ms, acquisition matrix: 80×80, resampled matrix: 128×128, FOV: 400 x 400 mm, slices: 46, slice thickness: 6 mm) sequences. The DWI protocol included diffusion weightings (*b*-values) of 0, 500, 1000, and 2000 s/mm^2^, with 1, 6, 6, and 12 diffusion directions, respectively. Each subject underwent this protocol at 5 different stations to produce a whole-body image. This protocol did not use respiratory gating.

In order to estimate the distortion due to *B*_*0*_ inhomogeneity using RPG, *b*=0 s/mm^2^ images were acquired in both the forward and reverse phase-encoded direction. The collection of the additional *b*=0 s/ mm^2^ image added 30 to 40 seconds to the protocol at each imaging station. The implementation details of the RPG algorithm has been described previously [5] and was applied using in-house software. Briefly, the forward and reverse images are smoothed and registered with one another using a least-squares cost function. This registration is used to calculate an initial estimate of the distortion. This procedure is then repeated several times, and, in each iteration, the smoothing is performed with a thinner kernel and the distortion estimate is refined. The result of this procedure is a 3D distortion map where each value in the volume represents the number of voxels it was displaced in the phase encoding direction due to *B*_*0*_ inhomogeneity. This distortion map can be applied to the raw diffusion images for distortion correction (DisCo). *B*_*0*_-related distortion is independent of diffusion weighting, so this map can be used to correct the entire diffusion dataset to produce post-DisCo images.

### Statistical Analysis

#### Anatomic Landmark Error Reduction Analysis

We first performed an anatomic landmark analysis to estimate the extent to which DisCo could reduce anatomic error in DWI of bone. The posterior edge of the vertebral column in the mid-sagittal plane was selected as the landmark because it is present in multiple stations and is discernable on both DWI (*b*=0) and *T*_*2*_-weighted images (the latter are less susceptible to distortion from *B*_*0*_ inhomogeneity). For each participant, we traced this landmark separately on the participant’s pre-DisCo *b*=0, post-DisCo *b*=0, and *T*_*2*_ images (for example, see Figure 1). Then, at each axial slice, we calculated the distance (along the anterior-posterior axis) between the pre-DisCo *b*=0 point annotation and the *T*_*2*_ point annotation as a measure of error. We further calculated the distance between the post-DisCo *b*=0 point annotation and the *T*_*2*_ point annotation in order to calculate the change in error after DisCo. We recorded the largest error reduction measured for each patient and provided summary statistics for this distribution. We also calculated the mean error (across the length of the spine) for each patient and compared these mean errors pre- and post-DisCo with a Wilcoxon signed-rank test.

**Figure 1.**
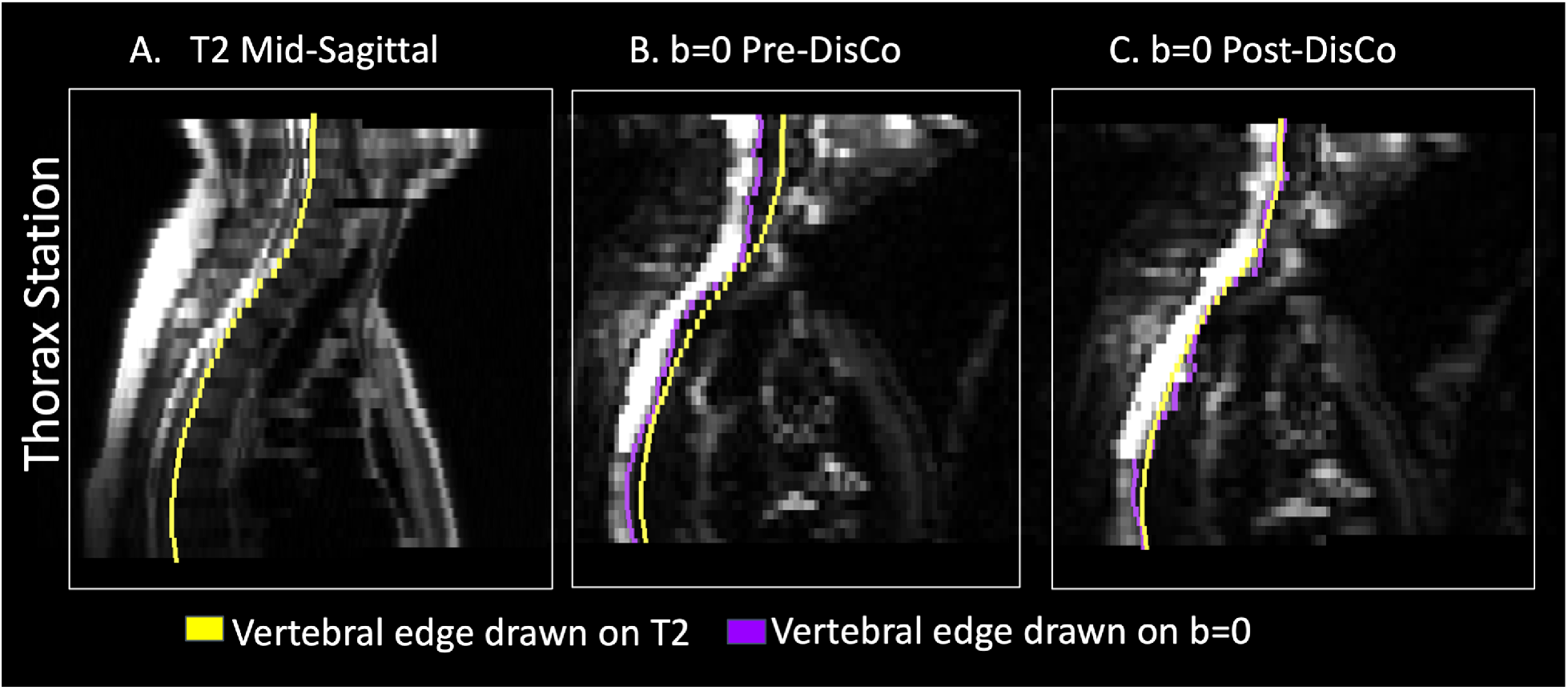
Example of tracing of posterior border of vertebrae for error change analysis. (A) Mid-sagittal slice of T2-weighted MRI from the Thorax Station for a single subject. The yellow represents the annotation of the posterior edge of the vertebral column on the T2. (B) Mid-sagittal slice of the DWI b=0 s/mm^2^ prior to DisCo. The purple in this panel represents the annotation of the posterior edge of the vertebral column on the pre-DisCo DWI b=0 s/mm^2^ (C) Mid-sagittal slice of DWI b=0 s/mm^2^ after DisCo. The purple in this panel represents the annotation of the posterior edge of the vertebral column on the post-DisCo DWI b=0 s/mm^2^. The average distance between the yellow and purple tracings (i.e., the error) decreases after application of DisCo. Abbreviations: DisCo=distortion correction.

#### Estimating Distortion of Bone Metastases

As discussed above, one practical application of DWI is its capability to detect bone metastases. Thus, we sought to characterize how distortions affected accurate localization of bone metastases in the present dataset. Bone metastases were identified based on available standard-of-care imaging (primarily CT, bone scan, and PET/CT) and DWI. These lesions were annotated manually on the diffusion images in MIM (MIM Software Inc, Cleveland, OH, USA) by a radiation oncologist (C.H.F.) with 4 years of experience. These annotations were then reviewed and confirmed by a fellowship-trained body radiologist (M.E.H.). Where applicable, the standard-of-care clinical imaging was also used to inform DWI lesion delineation. Only Stations 2, 3, and 4, were used in our analyses, as very few bone metastases were found in Stations 1 and 5. We refer to Stations 2, 3, and 4 as the Thorax, Abdomen, and Pelvis Station in the rest of this report.

For each lesion, we identified the horizontal slice in which the lesion was largest and selected it for analysis. Within that horizontal slice, a rectangular bounding box was drawn around each lesion, with the boundaries drawn 10 voxels from the lateral edge (in diffusion MRI space) of the lesion (for example, see Figure 2). The bounding box was then overlaid onto the distortion map. To quantify the extent to which distortion occurred within the lesion and in the immediate surrounding area, the root mean square (RMS) within the box was calculated using the following formula, as described previously [7]:

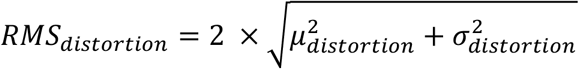

where *μ*_*distortion*_ and *σ*_*distortion*_ represent the mean and standard deviation of the distortion values of the voxels within the bounding box. We further generated 1000 bootstrap samples to obtain a 95% confidence interval for the mean RMS.

**Figure 2.**
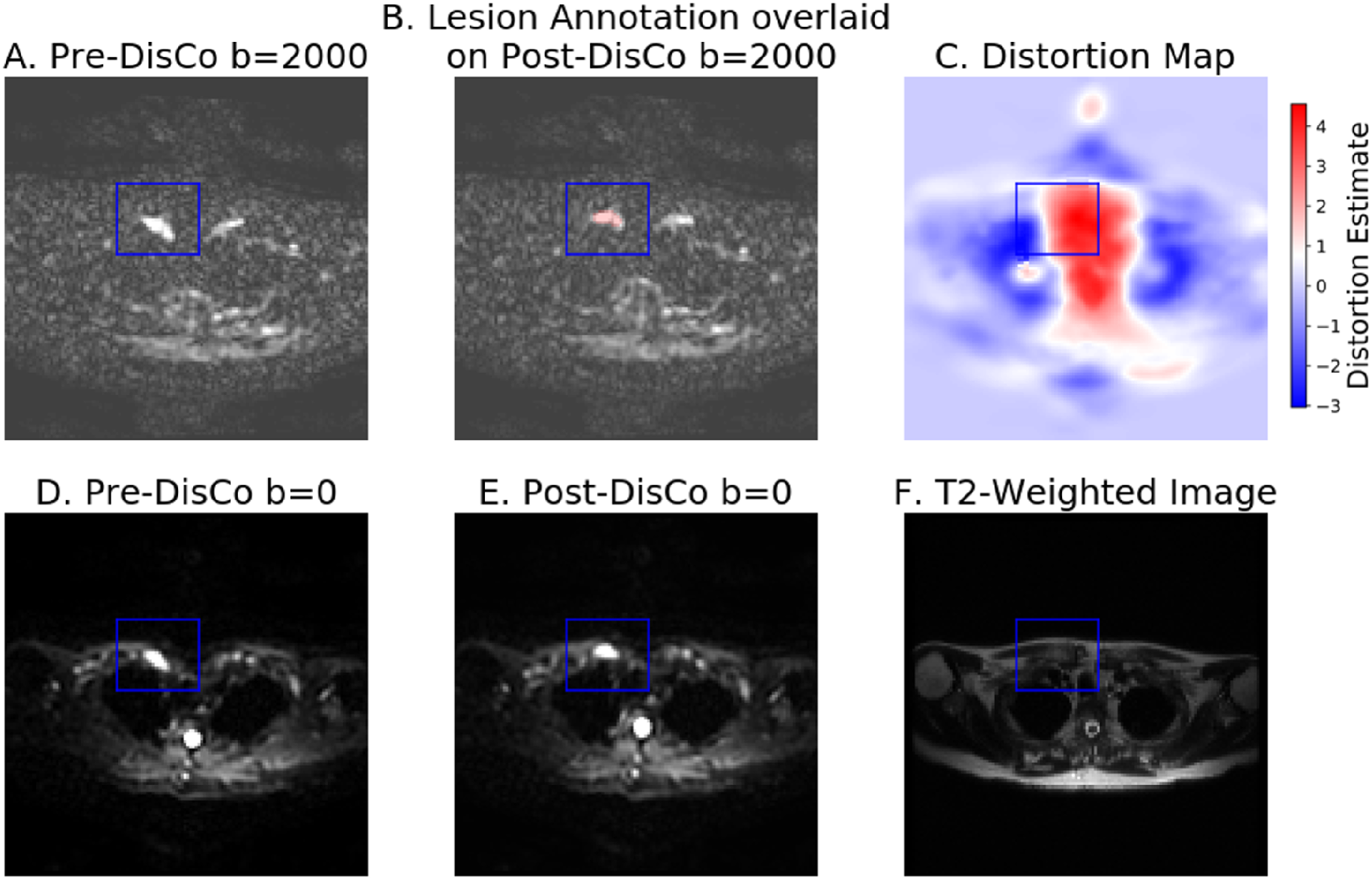
Example bone metastasis illustrating effect of B0 inhomogeneity induced distortion on DWI. In each subfigure, the slice corresponds to the horizontal slice at which the lesion is the largest (see Methods). The blue bounding box was drawn 10 voxels from the lateral edge of the lesion (in DWI space) in each of the 4 directions, then overlaid on the other images. A. Average b=2000 s/mm^2^ image prior to DisCo. B. Lesion annotation (pink) overlaid onto b=2000 s/mm^2^ image after DisCo. C. Estimation of the distortion within the slice. Voxel values represent the extent of displacement undergone by each voxel. Red and blue values denote displacement in the posterior and anterior direction, respectively. D. Average b=0 s/mm^2^ image prior to DisCo. E. Average b=0 s/mm^2^ image after DisCo. F. T2-weighted image at the same slice. Abbreviations: DisCo=distortion correction.

#### Assessing Similarity between Diffusion and Structural Images

The above analysis used DisCo to estimate the displacement of lesions in uncorrected images. Next, we conducted a mutual information analysis to assess the degree to which DisCo produces diffusion images that more faithfully reflect underlying anatomy of the *T*_*2*_-weighted images. The *T*_*2*_ image for each subject was resampled to match the resolution of the DWI images. For each lesion, we computed the normalized mutual information (MI) between the pre-DisCo *b*=0 image and *T*_*2*_ image, and the normalized MI between the post-DisCo *b*=0 image and *T*_*2*_ image. MI values were calculated for the entire horizontal slice [8]. We compared pre-DisCo and post-DisCo MI values with a Wilcoxon signed-rank test, with each lesion representing an observation. This Wilcoxon signed-rank analysis was then repeated for only the subset of lesions within each imaging station.

## Results

Our study included 127 participants with suspected bone metastases and complete whole-body multiparametric MRI data. 23 participants had visible bone metastases, with 75 individually annotated lesions across this subsample for inclusion in the bone lesion-level analysis.

### Anatomic Landmark Error Reduction Analysis

The mean (±SD) of largest error reduction (within each patient) was -11.8 mm (±3.6 mm). The distribution of largest error reduction for each patient is shown in Figure 3A. The largest observed error reduction was 29 mm, at a vertebral landmark in the thoracic vertebrae. Distortion correction led to consistent decreases in mean error for each subject (Wilcoxon signed-rank p<10^−20)^ (Figure 3).

**Figure 3.**
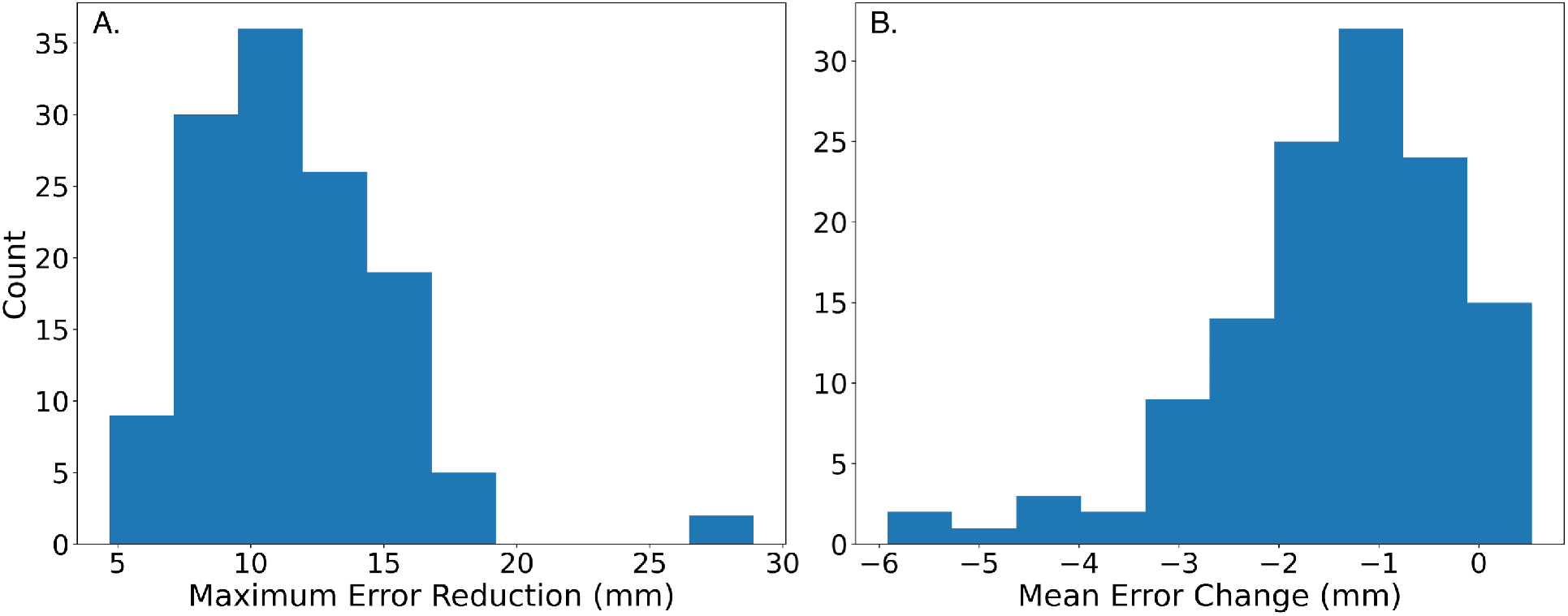
Application of DisCo leads to reduction in anatomic error of DWI. (A) Distribution of largest error reduction for each patient across our sample. (B) Mean change in error before and after application of DisCo. Abbreviations: DisCo = distortion correction.

### Estimating Distortion of Bone Metastases

The maximum RMS was 17.1 mm and was observed in a clavicular lesion in the Thorax Station. In the Abdomen and Pelvis Station, the maximum RMS values were 14.6 mm and 15.1 mm, respectively. Across all lesions, the average RMS was 6.0 mm (95% CI: 5.1-7.0). When grouped by station, the mean RMS for Thorax, Abdomen, and Pelvis Stations were 8.2 mm (95% CI: 6.6-9.8), 6.5 mm (95% CI: 4.5-8.6), and 4.1 mm (95% CI: 3.0-5.3), respectively. Examples of vertebral and femoral lesions are displayed in Figure 4. The distributions of RMS values are illustrated in Figure 5. RMS values did not significantly differ when using bounding boxes with edges at 7 or 12 voxels away from the lesion edges.

**Figure 4.**
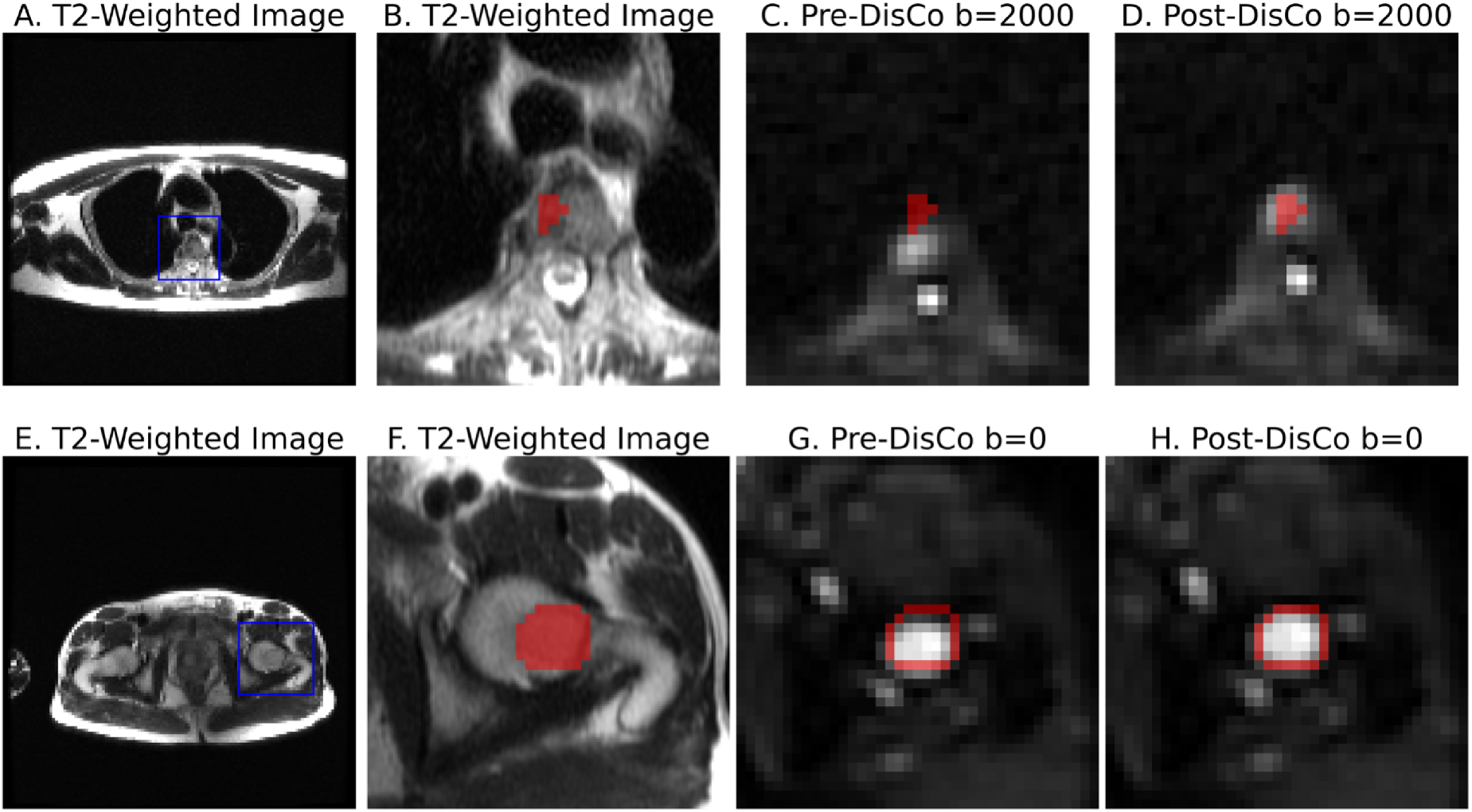
Examples of bone metastases as seen on b=0 s/mm^2^ DWI pre-and post-DisCo. The top row (A-D) shows a vertebral lesion in the Thorax Station. The bottom row (E-H) shows a lesion within the left femoral head in the Pelvis Station. (A,E) T2-weighted images at the corresponding level with blue bounding box drawn around lesion. (B, F) T2-weighted images zoomed in to visualize lesion and surrounding area. (C, G) Zoomed in DWI b=2000 s/mm^2^ and b=0 s/mm^2^ images before DisCo, respectively. (D, H) Zoomed in DWI b=0 s/mm^2^ images after DisCo. Outlines of the lesion annotations are overlaid in red. In both examples, the lesions were translated anteriorly following DisCo. Without DisCo, lesion localization may have erred posteriorly. Abbreviations: DisCo=distortion correction.

**Figure 5.**
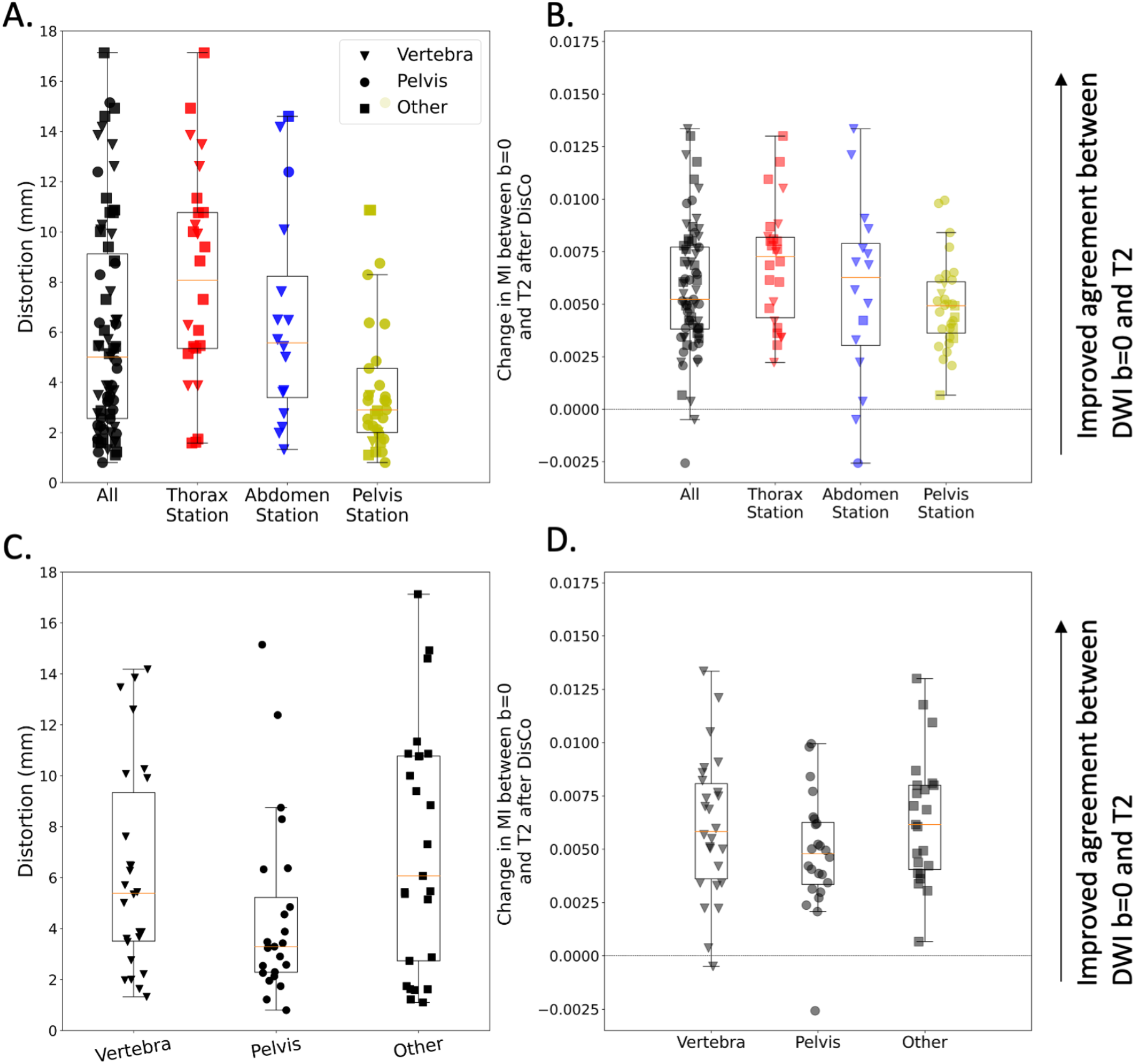
Distribution of RMS of distortion and MI between b=0 s/mm^*2*^ and T2-weighted images. (A) Distortion for lesions across all stations (black) as well as the distribution within specific imaging Stations (red, blue, and yellow). Distortion was estimated by calculating the RMS of the values in the distortion map that correspond to the lesion annotation and immediately surrounding areas (see Methods). (B) Change in MI values between the DWI b=0 s/mm^2^ and T2-weighted images after DisCo. A value larger than 0 indicates improved agreement between DWI b=0 and T2-weighted images. Again, we plot the distribution for all lesions as well as the breakdown for different Stations. (C) Distortion for lesions broken down by anatomic group. (D) Change in MI values between the DWI b=0 s/mm^2^ and T2-weighted images after DisCo broken down by anatomic group. DisCo led to consistent increases in similarity between DWI b=0 s/mm^2^ and T2-weighted images. Abbreviations: RMS=root mean square, MI=mutual information.

### Assessing Similarity between Diffusion and Structural Images

Applying DisCo led to consistently increased MI between the DWI *b*=0 s/mm^2^ and *T*_*2*_ images. When comparing pre-DisCo to post-DisCo MI values with Wilcoxon signed-rank tests, we found that the post-DisCo MI values were significantly higher (p<10^−12^), indicating the RPG distortion correction improved similarity of DWI with the anatomic *T*_*2*_ images (Figure 5). This was consistent for each of the imaging stations analyzed (Thorax Station: p<10^−5^, Abdomen Station: p=0.0004, Pelvis Station: p<10^−5^).

## Discussion

Application of RPG to DWI led to consistent improvement in the anatomic accuracy of DWI of the vertebral skeleton. We found reductions in anatomic inaccuracy of up to 29 mm using RPG. On average, the participants’ largest error reduction was 11.8 mm (±3.6). Similarly, for bone metastases, we observed a mean displacement of 6.0 mm (95% CI: 5.1-7.0) and displacement of up to 17 mm, which are meaningful distortions in potential clinical applications like image-guided biopsy (where sampling the more hypercellular part of the lesion may be desired) or image-guided treatments like stereotactic radiotherapy [9]. As DWI continues to take on an increasing role in musculoskeletal imaging [1], distortion correction may facilitate improved localization of bone tissue and associated pathology, especially in applications where high precision is required.

Prior studies have demonstrated that *B*_*0*_ inhomogeneities affect organ or lesion localization in the prostate and breast [7, 8, 10]. The extent of *B*_*0*_ inhomogeneities and their consequent artifacts depend, in part, on the anatomic environment of a region of interest or lesion (e.g., how close the lesion is to an air-tissue interface). Since bone occupies anatomic environments distinct from those of prostate and breast, it remains to be determined whether and to what extent these artifacts affect DWI of bone. Our findings here suggest that, like prostate and breast, bone is also susceptible to meaningful distortion.

Estimated distortion of lesions from *B*_*0*_ inhomogeneity was greatest in the Thorax Station and smallest in the Pelvis Station. The elevated RMS values in the Thorax Station, relative to the other imaging stations, may be due to a number of factors. As noted above, the magnitude of *B*_*0*_ inhomogeneities and their consequent artifacts depend partly on proximity to an air-tissue interface. The Thorax Station images several bones close to air-tissue interfaces, including the clavicles, ribs, and scapulae. Since these bones are all close to air-tissue interfaces, there are several places within the FOV of the Thorax Station where bone and bone metastases may be prone to heavy distortion. However, the clavicles, ribs, and sternum are also subject to respiratory motion, and we are not able to separate these effects in this study. Future studies might benefit from controlling for respiratory motion, such as abdominal compression, active breathing control, or respiratory gating [11]. Nevertheless, we found that the vertebrae of the Thorax Station, which are less affected by respiratory motion, still exhibited elevated RMS suggesting that not all of the distortion observed in the Thorax Station in our study can be explained by respiratory motion.

In addition to the spatial displacements that we have demonstrated in the skeletal landmark and bone lesion analyses, *B*_*0*_ inhomogeneities can also lead to geometric distortions. As demonstrated in Figure 2C, voxels undergo varying levels of displacement along the phase-encoding gradient. Fluctuating amounts of distortion within and around bone can lead to contraction, expansion, or other geometric distortion of the bone tissue. The metastasis in the left femoral head shown in Figure 4E-H underwent *B*_*0*_ inhomogeneity induced contraction, as can be appreciated with the dark strip of voxels at the anterior edge of the lesion in Figure 4G. The right clavicular lesion in Figure 2 also clearly underwent geometric distortion. One clinical application where geometric distortion may cause concern is in the tracking of treatment response of bone metastases, where DWI has demonstrated utility [12, 13]. Artificial geometric distortions from *B*_*0*_ inhomogeneity like those demonstrated in Figures 2 and 4 could lead to misinterpretations in patients’ responses to therapy, such as mistaking imaging-related contraction or expansion distortions for tumor shrinkage or growth.

There are several tools available to correct for these artifacts and reduce the likelihood of missing a target or misinterpreting treatment response [4–6]. In the RPG method used here, a correction that can be applied to the diffusion dataset is simultaneously generated while calculating the voxel-wise displacements; this method was chosen for its efficiency, as little additional scan time (30-40 seconds per station) was required. A salient question, however, is whether this correction actually produces images that more accurately reflect anatomy. We demonstrated consistent reductions in mean error across subjects, as well as the increases in MI between DWI *b*=0 and *T*_*2*_ images after correcting for distortion. This is in line with prior work that demonstrated improved similarity of DWI with *T*_*2*_ anatomic imaging after RPG [8, 10]. Inspection of data revealed some instances of increases in measured error after the application of RPG. These may be due to actual movement of the patient or internal organs (e.g., via breathing or peristalsis) between DWI and *T*_*2*_ acquisitions. Nevertheless, the net effect of RPG was to consistently reduce measured errors. Taken together, our findings suggest that the RPG method is capable of generating diffusion images that more closely represent the true anatomy.

Limitations of our study include the potential effects of respiration on our distortion estimates that were discussed above. Beyond respiratory motion alone, slight differences in respiratory volume could also lead to artifacts. Thus, even if image acquisition were gated with the respiratory cycle, it is likely that some residual distortion would remain. Our group is actively working on imaging protocols and processing methods to address this residual distortion, which is relevant not only to imaging of bones near the lungs, but also to imaging of other organs near the diaphragm, such as the liver and pancreas. We also note that our whole-body imaging protocol does not capture the bones of the distal extremities. In the case of cancer, metastases in the distal extremities are uncommon. In other settings of musculoskeletal DWI, however, such as knee imaging, these locations are of interest. A larger-bore system might allow inclusion within the field of view. Future studies dedicated to characterizing the distortion in these anatomic locations will be needed as the artifacts will likely exhibit different characteristics. A final limitation is that our study sample only consisted of prostate cancer patients. Bone is also a common site of metastases in other cancers, particularly lung and breast cancer. DWI has been successfully used to visualize bone metastases in other cancers (for breast see [12]; for melanoma, see [14]). Lesion appearance may vary on DWI by cancer type, but there is no *a priori* reason to suspect that distortion effects would be substantially affected by cancer histology.

In summary, we found that *B*_*0*_ inhomogeneity results in distortion of whole-body diffusion images that leads to artifactual displacement of bone and bone metastases. These distortions may be severe enough to interfere with accurate biopsy or stereotactic treatment. Moreover, these distortions could complicate interpretation of tumor shrinkage or growth. The RPG technique used here is an efficient solution (costing only 30-40 seconds per station) for reducing distortion artifacts in whole-body DWI.

## Data Availability

For inquiries about the code and data used in this study, please contact the corresponding author.

## Abbreviations

MI: Mutual information
RMS: Root mean square
RPG: Reverse polarity gradient

